# Self-sampling of capillary blood for serological testing of SARS-CoV-2 by COVID-19 IgG ELISA

**DOI:** 10.1101/2020.09.25.20183459

**Authors:** Lottie Brown, Rachel L Byrne, Alice Fraser, Sophie I Owen, Ana I Cubas-Atienzar, Chris Williams, Grant A Kay, Luis E Cuevas, Joseph R A Fitchett, Tom Fletcher, Gala Garrod, Konstantina Kontogianni, Sanjeev Krishna, Stefanie Menzies, Tim Planche, Chris Sainter, Henry M Staines, Lance Turtle, Emily R Adams

## Abstract

Serological testing is emerging as a powerful tool to progress our understanding of COVID-19 exposure, transmission and immune response. Large-scale testing is limited by the need for in-person blood collection by staff trained in venepuncture. Capillary blood self-sampling and postage to laboratories for analysis could provide a reliable alternative.

Two-hundred and nine matched venous and capillary blood samples were obtained from thirty nine participants and analysed using a COVID-19 IgG ELISA to detect antibodies against SARS-CoV-2.

Thirty seven out of thirty eight participants were able to self-collect an adequate sample of capillary blood (≥50 μl). Using plasma from venous blood collected in lithium heparin as the reference standard, matched capillary blood samples, collected in lithium heparin-treated tubes and on filter paper as dried blood spots, achieved a Cohen’s kappa coefficient of >0.88 (near-perfect agreement). Storage of capillary blood at room temperature for up to 7 days post sampling did not affect concordance.

Our results indicate that capillary blood self-sampling is a reliable and feasible alternative to venepuncture for serological assessment in COVID-19.

## Introduction

Serological testing is emerging as a powerful tool to progress our understanding of SARS-CoV-2 transmission. Serological tests are able to detect specific antibodies for severe acute respiratory syndrome coronavirus 2 (SARS-CoV-2), the causative agent of COVID-19. In individuals who have been infected with the virus, immunoglobulin G (IgG) antibodies specific to viral proteins appear approximately 6-15 days after infection (Zhao *et al*., 2020). The longevity of this antibody response and whether it confers neutralising or protective immunity is still uncertain, but evidence suggests IgG antibodies may be detected for months following infection (Liu *et al*., 2020; Reifer *et al*., 2020; Sekine *et al*., 2020; Staines *et al*., 2020; Wajnberg *et al*., 2020; Wu *et al*., 2020). Wide-scale serological testing is key to improving our understanding of longitudinal antibody responses and immunity in COVID-19 and for epidemiological analyses.

Many of the currently available antibody tests for SARS-CoV-2, such as those that use enzyme-linked immunosorbent assay (ELISA) technology, require serum or plasma collected by venepuncture. Collection of venous blood is labour and resource-intensive, as it demands trained healthcare staff, facilities, personal protective equipment (PPE), and that the participant must attend a health facility in person for collection. Bringing individuals into the healthcare environment for sampling increases the risk of COVID-19 transmission (S.C.Y. *et al*., 2020; Sikkema *et al*., 2020). On a large-scale, this type of sampling is impractical, particularly in low and middle-income countries (LMICs), where resources and healthcare personnel are limited.

An alternative is surveillance of antibody response by rapid lateral flow immunoassays (LFIAs). However, the performance of the first generation of assays is variable, even when conducted by trained staff in a clinical setting (Whitman *et al*., 2020). A U.K. seroprevalence study evaluated finger prick self-testing by LFIAs at home and found highly variable sensitivity (21% to 92%), that was significantly inferior to ELISAs performed on venous blood (Flower *et al*., 2020). Studies have shown high sensitivity of ELISAs for the detection of SARS-CoV-2 antibodies in venous blood (Adams *et al*., 2020; Lisboa Bastos *et al*., 2020; Staines *et al*., 2020). Capillary blood is a well-established specimen in ELISA diagnostics but it is yet to be evaluated in SARS-CoV-2 infection. If individuals were able to self-sample blood in their own home and post to a reference laboratory, this would mitigate the potential transmission risks for inpatient sampling, boost community surveillance and open sampling to a wider population; yet retain the use of more sensitive ELISAs.

The aims of this study were to determine the feasibility of capillary blood self-sampling and to compare the COVID-19 IgG ELISA results from samples derived from capillary blood with those from blood samples obtained by venepuncture. We investigated the stability of SARS-CoV-2 antibodies in capillary blood stored at room temperature over 7 days. Additionally, we determined whether the performance of COVID-19 IgG ELISA was equivalent on capillary blood collected on dried blood spots (DBS).

## Methods

### Study participants

Individuals were recruited based on past positive SARS-CoV-2 PCR result (n = 8) and/or positive antibody test (n = 11) with strong clinical suspicion of COVID-19. Healthy volunteers who had not experienced symptoms of COVID-19 were enrolled as negative controls. Participants were identified from an existing study (the Flavimmune study, 16/NW/0160), after ethical approval was sought for repurposing the study for COVID-19. Participants gave consent for the use of their samples for this purpose. A total of 18 participants were classified as serologically positive and 21 as serologically negative based on the results of COVID-19 IgG ELISA performed plasma from venous blood collected in lithium heparin tubes, which was taken as a reference.

### Venous blood collection

Healthcare staff trained in venepuncture collected four tubes (approximately 20 ml) of venous blood per participant. Blood collection tubes (BD Vacutainer®, UK); one treated with lithium heparin, one with potassium ethylenediaminetetraacetic acid (EDTA) and one with sodium citrate were used for plasma separation. A final tube, treated with silica additive, was used for serum separation. Blood was processed following the manufacturer’s instructions according to tube type.

### Capillary blood collection

Capillary blood was collected using Microvette® 100 capillary tubes by Sarstedt® (Sarstedt®, Nümbrecht, Germany), which are treated with lithium heparin. The self-sampling procedure was explained to each participant by the investigator. Participants were advised to place their non-dominant hand in warm water for 1-3 minutes, before thoroughly drying with clean paper towels. Participants then cleaned the ring finger with an alcohol wipe and punctured with a 2.8 mm SurgiLance® lancet (Medipurpose, New Malden, Surrey, UK). Blood flow was encouraged by massaging the finger in the direction of the puncture site until a large droplet of blood formed. The participant then brought the capillary tube in contact with the blood to draw it into the tube and continue this process until approximately 50 μl of blood had been collected. An additional four capillary samples were taken by the study investigators, dependent upon sufficient blood flow and absence of clotting. A maximum of four puncture sites, each on separate fingers, were used for collection. The volume of self-collected capillary blood was recorded.

### Sample storage and processing

The first self-collected capillary sample was designated “day 1 refrigerated”, and immediately stored at 2-8°C along with venous plasma and serum. The remaining capillary blood samples were stored at room temperature (21 – 25°C) for testing at days 1, 3, 5 and 7. Immediately before testing, capillary blood samples were centrifuged for 5 minutes at 10,000 x g and 25°C to obtain plasma.

### Detection of SARS-CoV-2 antibodies in capillary and venous blood

On the day following sample collection (day 1), the venous and capillary samples (refrigerated and stored at room temperate) were run in triplicate on the COVID-19 IgG ELISA (Omega Diagnostics Ltd, Littleport, Cambridgeshire, UK) according to manufacturer’s instructions; positive, cut-off, and negative controls were run in duplicate. Samples with a mean OD value of 10% greater than the cut-off control, as defined in the manufacturer’s instructions, were regarded as positive for SARS-CoV-2 antibodies (Adams *et al*., 2020; Staines *et al*., 2020). Participants were deemed positive or negative based on their day 1 venous plasma (lithium heparin) IgG result. On day 1, 3, 5 and 7, plasma was obtained from stored capillary blood samples (room temperature) and run in triplicate on the COVID-19 IgG ELISA. If fewer than five capillary samples had been obtained, priority was given to testing on day 5, day 1, day 3 and then day 7.

### Dried blood spot (DBS)

To determine whether comparable results were achieved using capillary blood deposited on dried blood spots (DBS), 50 μl of capillary blood stored in lithium-heparin treated tubes at room temperature for 5 days was mixed by vortex, pipetted onto 903 Whatman card and left to dry (Cytiva, Malborough, USA). After one hour, a single 6 mm disc was punched out and eluted in 1000 μl of COVID-19 IgG ELISA diluent and incubated at 2-8°C for 16 hours. The eluate was then treated as a diluted sample and run in triplicate in the ELISA.

### Statistical analysis

The mean absorbance (OD_450nm_) values for each sample and control were tabulated. Agreement of COVID-19 IgG ELISA results on different blood samples was assessed by determining the Cohen’s kappa coefficient with 95% confidence intervals (95% CI). Raw OD values were normalised to allow comparison across ELISA plates. For each plate the mean cut-off OD value plus 10% was subtracted from each mean sample OD value. The resulting value was divided by the mean positive control OD value to give a normalised OD. Statistical analyses were performed using Prism (version 8, GraphPad, USA). Correlations between OD values were assessed using Spearman’s rank test and a p value <0.05 was considered statistically significant. Agreement in normalised OD results across sample types was measured by Bland-Altman mean difference.

## Results

### Participant characteristics

A total of 39 participants were included, of which 22 (56.4%) were female (Table 1). The median age was 37 years (range = 23-64 years). Only one participant was not able to self-collect an adequate capillary blood sample ≥ 50 μl.

**Table 1.**
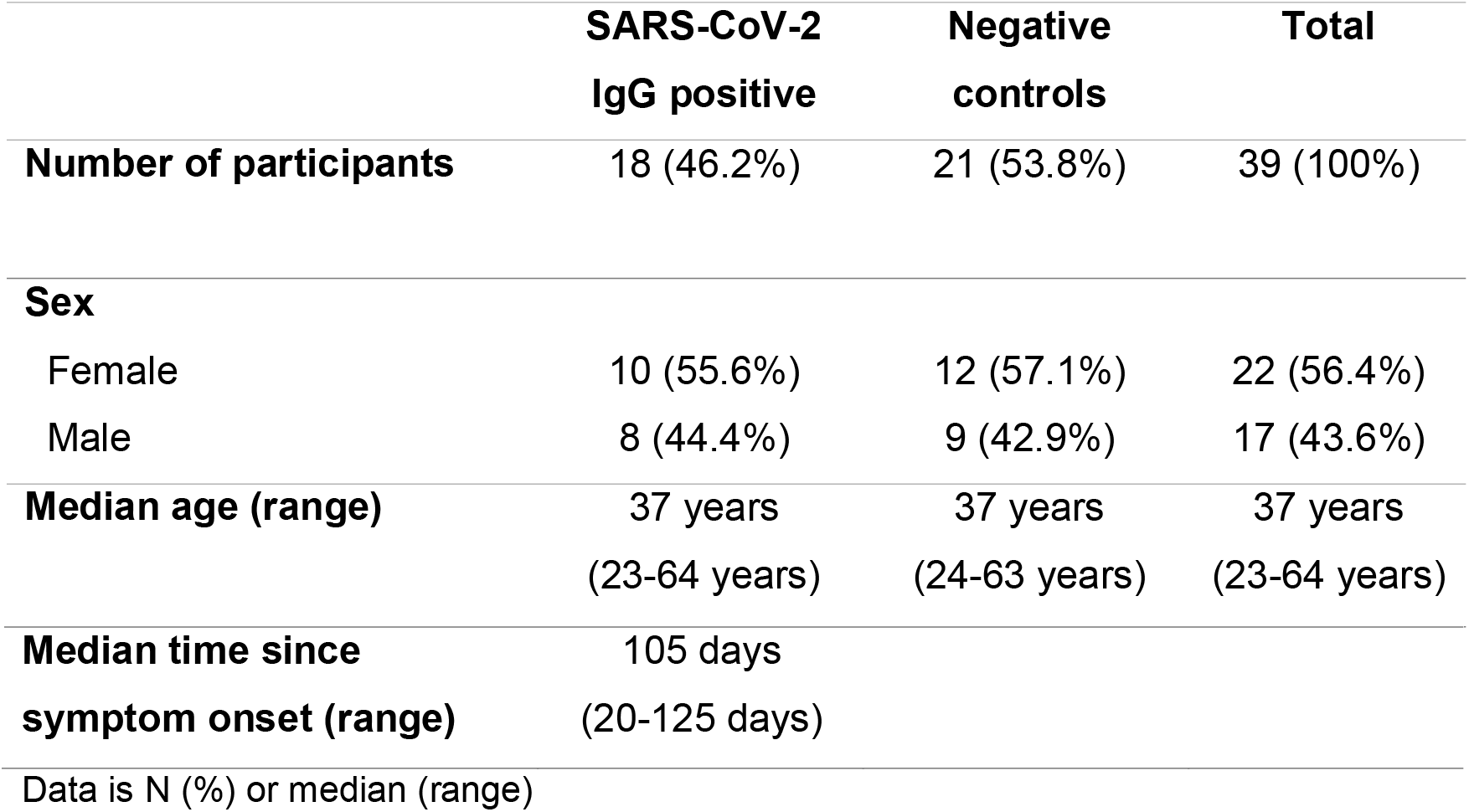
Participant demographics.

### Agreement between COVID-19 IgG ELISA results, using capillary and venous blood samples

Here, we report high agreement between capillary and venous blood samples for the detection of SARS-CoV-2 antibodies by IgG ELISA. As shown in Table 2, agreement of the samples was consistently high (≥94%) and storage at room temperature for 7 days did not affect concordance. Discordance occurred between only five of 209 matched venous and capillary samples (2.4%), giving Cohen’s kappa coefficient >0.88 for all sample types (near-perfect agreement). The cases of disagreement only occurred in samples with OD values close to the cut-off, when the reference venous blood was positive and capillary blood was indeterminate (Adams *et al*., 2020; Staines *et al*., 2020).

**Table 2.**
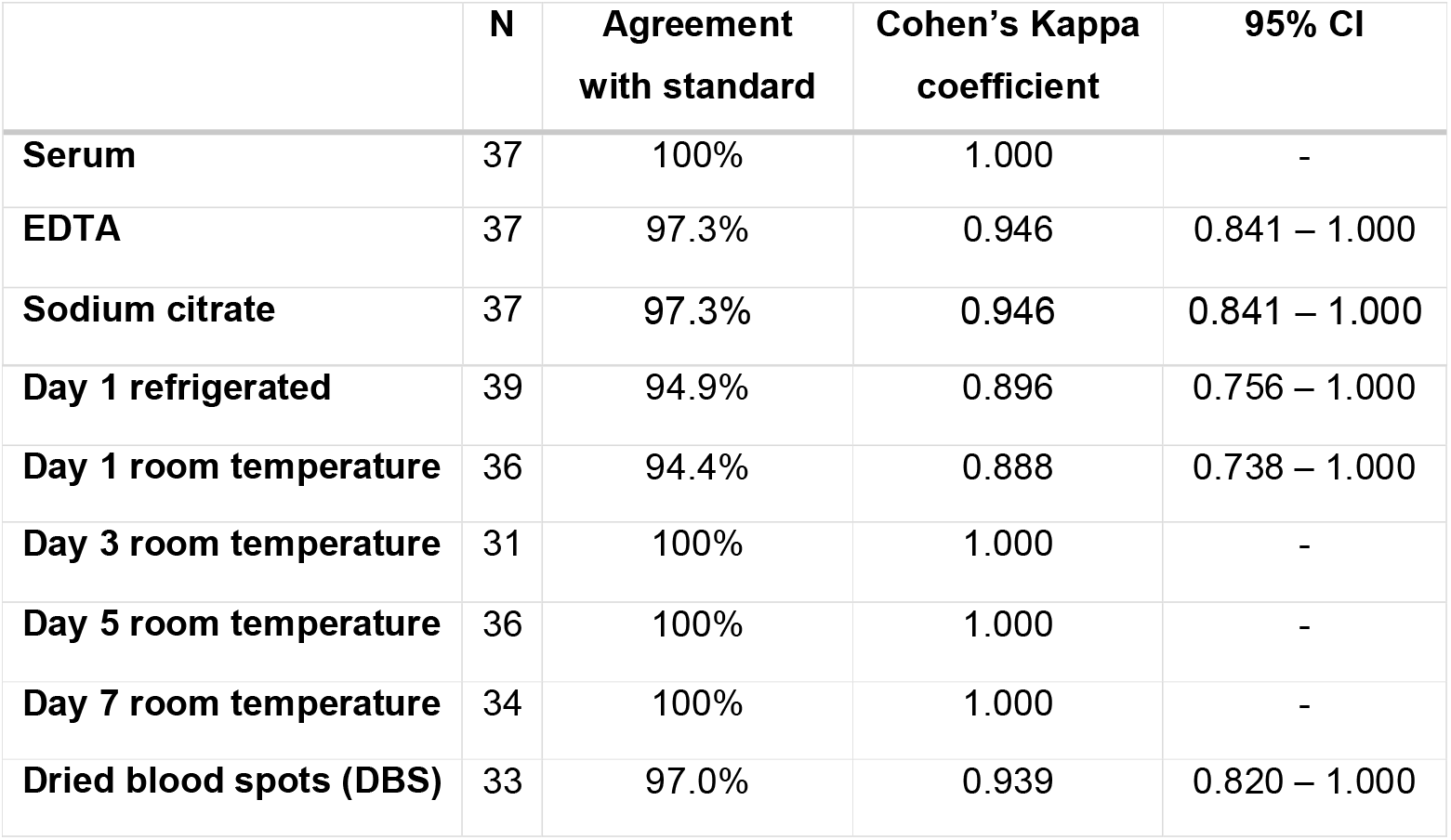
Agreement between results of the COVID-19 IgG ELISA performed on different sample types and days post sampling, compared with a reference standard of plasma from venous blood collected in lithium heparin-treated tubes.

The OD detected by ELISA for 209 matched venous and capillary blood samples were standardised and plotted. We found a strong correlation between the matched OD values (r = 0.92, 95% CI 0.90-0.94, p<0.0001, Figure 1) and minimal differences in results across all sample types (Bland-Altman Bias −0.0028 ± 0.19).

**Figure 1:**
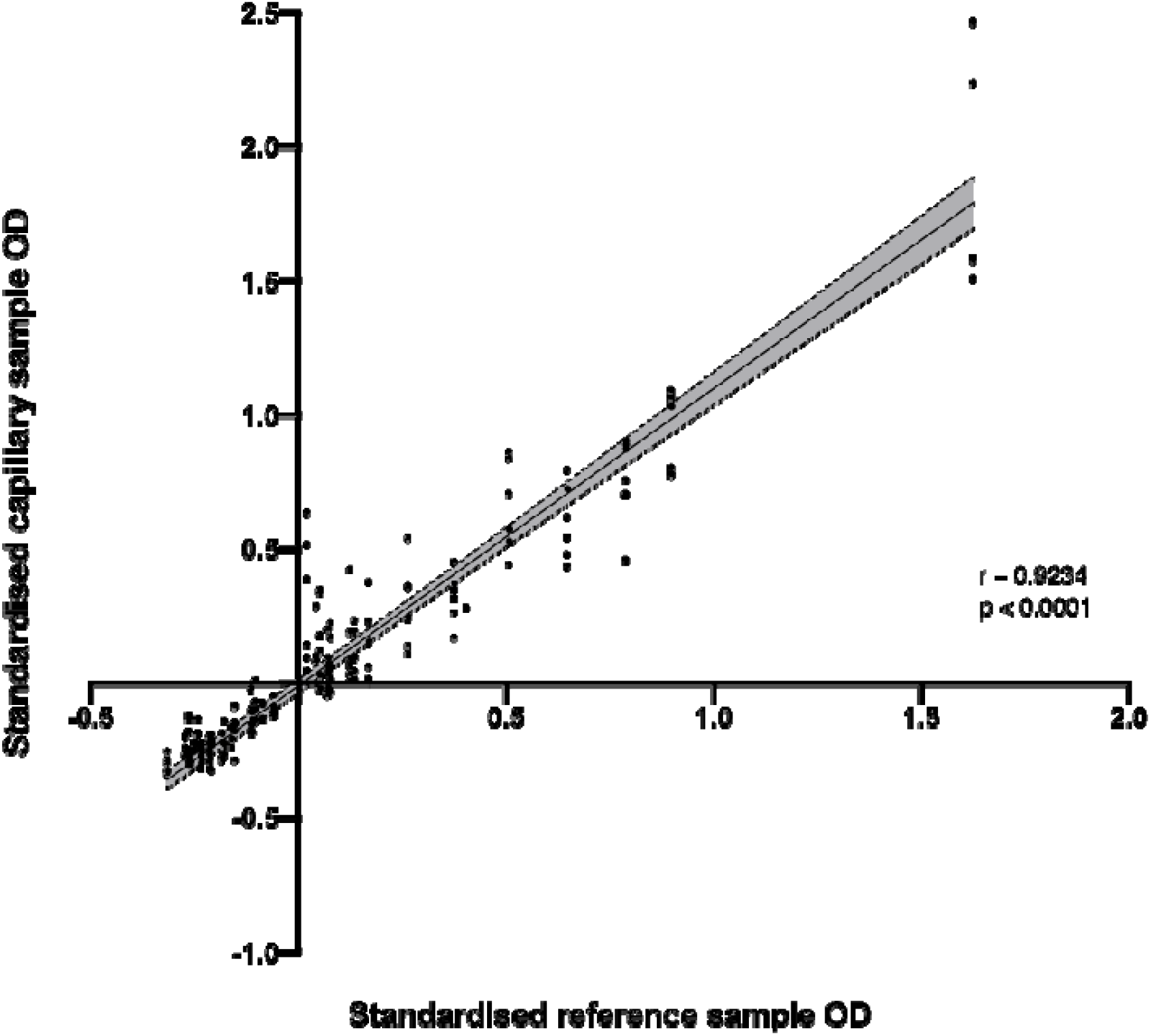
Linear relationship between standardised OD values of matched venous plasma collected in lithium heparin treated tubes and all capillary samples.

This figure displays the relationship between the standardised OD values of venous plasma (collected in lithium heparin treated tubes) and of capillary blood samples (collected on all days in lithium heparin-treated tubes and deposited onto DBS). The black line indicates the linear regression result and shaded area is 95% confidence interval.

## Discussion

Here, we report that results from a COVID-19 IgG ELISA performed on samples derived from capillary blood samples, which can be self-collected, are comparable to blood taken by venepuncture for the detection of SARS-CoV-2-specific antibodies. One of the bottlenecks to antibody testing in the COVID-19 pandemic, is the need for in-person venous blood collection by healthcare staff. The utilisation of an easy to self-collect specimen such as capillary blood offers improved access to serological testing. Almost all participants in our study (97.4%) were able to self-collect an adequate capillary blood sample for the Omega SARS-CoV-2 assay, which indicates self-sampling is a feasible alternative to venepuncture. In addition, we show SARS-CoV-2 antibodies within capillary blood are stable for up to one week (the limit of testing) at room temperature (21 - 25°C). This mitigates the need for cold chain transportation and reduces the pressure on the postal service and laboratory staff for immediate testing.

We report a high level of agreement between venous and capillary blood samples with only 2.4% of discordant samples across the study. Furthermore, we found that the OD values of matched venous plasma and capillary samples were highly similar. Type of capillary blood (in lithium heparin-treated capillary tubes or deposited onto filter paper as DBS) did not affect equivalency. Capillary blood stored in lithium heparin-treated tubes can be processed immediately on receipt of samples. There are more manual pre-analytical steps when processing DBS samples, as they must be punched out and eluted before ELISA which can be time-consuming. An advantage of DBS samples over capillary blood tubes is the minimal biohazard risk during transport. Unlike liquid blood samples, DBS do not require special handling during postage or transport. Antibodies are known to remain stable in DBS for weeks at room temperature, although this has not yet been validated in antibodies against SARS-CoV-2.

There was disagreement between the results of venous and capillary blood sampling in only 5 (2.4%) of samples in our study. In every case of disagreement, the venous sample was low level positive, and the capillary sample was indeterminate. Therefore, if the results of the test are of particular importance to an individual, an indeterminate test could be repeated with a venous blood sample. As only 5 (2.9%) capillary samples gave indeterminate results, this still has the potential to reduce the amount of sampling necessary greatly.

Three studies have evaluated the use of DBS in comparison to venous blood samples for SARS-CoV-2 serology and found high concordance (Karp *et al*., 2020; McDade *et al*., 2020; Morley *et al*., 2020). In this study, the method of collection presented little difficulty for our volunteers, with only one person not able to collect an adequate sample. However, more research exploring capillary blood collection preferences and acceptability in target populations is needed.

We plan to validate this method in future studies of large patient cohorts having contributed this proof of concept and completed the pilot. Both methods of capillary blood sampling are simple and inexpensive and are suited to LMICs, particularly where there is a lack of trained healthcare staff or geographically dispersed populations. Our findings indicate antibodies for SARS-CoV-2 are stable in capillary blood stored at 21 - 25°C. However, to confirm the suitability of this method for tropical LMICs, we will repeat our study after storing capillary blood samples (collected in capillary tubes and as DBS) at a higher temperature.

Our results show that capillary blood self-sampling is a reliable and feasible alternative to venepuncture for serological assessment in COVID-19. This method will be particularly useful in large-scale disease surveillance and longitudinal research of immune response in COVID-19.

## Data Availability

Raw data were generated at Liverpool School of Tropical Medicine. Derived data supporting the findings of this study are available from the corresponding author ERA on request.

## Acknowledgements

We are grateful to all the volunteers who enrolled into our study, for dedicating time during the pandemic to be involved in research.

## Author contributions

ERA, JRAF, LEC and CS conceived and designed the study. LT, HM, SK and TP obtained ethical approval. LB, RLB, SIO, AICA, GK, TF, SM, GG and KK collected clinical data and patient samples. LB and CW obtained the laboratory data. LB, AF, SIO, CW analysed the laboratory data. LB wrote the first draft of the manuscript. All authors reviewed and approved the final version of the manuscript for submission.

## Ethics

The study was reviewed and approved by the National Health Service Research Ethics Committee Liverpool Central IRAS number 16/NW/0160.

## References

Adams, E. R. et al. (2020) ‘Rapid development of COVID-19 rapid diagnostics for low resource settings: accelerating delivery through transparency, responsiveness, and open collaboration’, medRxiv. doi: 10.1101/2020.04.29.20082099.

Flower, B. et al. (2020) ‘Clinical and laboratory evaluation of SARS-CoV-2 lateral flow assays for use in a national COVID-19 seroprevalence survey.’, Thorax. BMJ Publishing Group Ltd. doi: 10.1136/thoraxjnl-2020-215732.

Karp, D. G. et al. (2020) ‘A serological assay to detect SARS-CoV-2 antibodies in at-home collected finger-prick dried blood spots.’, *medRxiv L: the preprint server for health sciences*. doi: 10.1101/2020.05.29.20116004.

Lisboa Bastos, M. et al. (2020) ‘Diagnostic accuracy of serological tests for covid-19: systematic review and meta-analysis’, BMJ. doi: 10.1136/bmj.m2516.

Liu, A. et al. (2020) ‘Antibody responses against SARS-CoV-2 in COVID-19 patients’, Journal of Medical Virology. doi: 10.1002/jmv.26241.

McDade, T. W. et al. (2020) ‘High seroprevalence for SARS-CoV-2 among household members of essential workers detected using a dried blood spot assay’, medRxiv. doi: 10.1101/2020.06.01.20119602.

Morley, G. L. et al. (2020) ‘Sensitive detection of SARS-CoV-2-specific-antibodies in dried blood spot samples’, medRxiv. doi: 10.1101/2020.07.01.20144295.

Reifer, J. et al. (2020) ‘SARS-CoV-2 IgG Antibody Responses in New York City’, medRxiv. Cold Spring Harbor Laboratory Press, p. 2020.05.23.20111427. doi: 10.1101/2020.05.23.20111427.

S.C.Y., W. et al. (2020) ‘Risk of nosocomial transmission of coronavirus disease 2019: an experience in a general ward setting in Hong Kong’, Journal of Hospital Infection.

Sekine, T. et al. (2020) ‘Robust T cell immunity in convalescent individuals with asymptomatic or mild COVID-19’, *bioRxiv*. doi: 10.1101/2020.06.29.174888.

Sikkema, R. S. et al. (2020) ‘COVID-19 in health-care workers in three hospitals in the south of the Netherlands: a cross-sectional study’, The Lancet Infectious Diseases. doi: 10.1016/S1473-3099(20)30527-2.

Staines, H. M. et al. (2020) ‘Dynamics of IgG seroconversion and pathophysiology of COVID-19 infections’, medRxiv. doi: 10.1101/2020.06.07.20124636.

Wajnberg, A. et al. (2020) ‘Title: SARS-CoV-2 infection induces robust, neutralizing antibody responses that are 1 stable for at least three months 2 3’, medRxiv. doi: 10.1101/2020.07.14.20151126.

Whitman, J. D. et al. (2020) ‘Test performance evaluation of SARS-CoV-2 serological assays’, medRxiv. doi: 10.1101/2020.04.25.20074856.

Wu, J. et al. (2020) ‘SARS-CoV-2 infection induces sustained humoral immune responses in convalescent patients following symptomatic COVID-19 Correspondence’, medRxiv. doi: 10.1101/2020.07.21.20159178.

Zhao, J. et al. (2020) ‘Antibody responses to SARS-CoV-2 in patients of novel coronavirus disease 2019’, Clinical infectious diseases: an official publication of the Infectious Diseases Society of America. doi: 10.1093/cid/ciaa344.

